# Spatial Patterns of White Matter Hyperintensities: A Systematic Review

**DOI:** 10.1101/2023.02.13.23285878

**Authors:** Jonas Botz, Valerie Lohner, Markus D. Schirmer

## Abstract

**Background:** White matter hyperintensities are an important marker of cerebral small vessel disease. This disease burden is commonly described as hyperintense areas in the cerebral white matter, as seen on T2-weighted fluid attenuated inversion recovery magnetic resonance imaging data. Studies have demonstrated associations with various cognitive impairments, neurological diseases, and neuropathologies, as well as clinical and risk factors, such as age, sex, and hypertension. Due to their heterogeneous appearance in location and size, studies have started to investigate spatial distributions and patterns, beyond summarizing this cerebrovascular disease burden in a single metric - its volume. Here, we review the evidence of association of white matter hyperintensity spatial patterns with its risk factors and clinical diagnoses.

**Design/Methods:** We performed a systematic review in accordance with the Preferred Reporting Items for Systematic Reviews and Meta-Analysis (PRISMA) Statement. We used the standards for reporting vascular changes on neuroimaging criteria to construct a search string for literature search on PubMed. Studies written in English from the earliest records available until January 31st, 2023, were eligible for inclusion if they reported on spatial patterns of white matter hyperintensities of presumed vascular origin.

**Results:** A total of 380 studies were identified by the initial literature search, of which 41 studies satisfied the inclusion criteria. These studies included cohorts based on mild cognitive impairment (15/41), Alzheimer’s Disease (14/41), Dementia (5/41), Parkinson’s Disease (3/41), and subjective cognitive decline (2/41). Additionally, 6 of 41 studies investigated cognitively normal, older cohorts, two of which were population-based, or other clinical findings such as acute ischemic stroke or reduced cardiac output. Cohorts ranged from 32 to 882 patients/participants (median cohort size 191.5 and 51.6 % female (range: 17.9 - 81.3 %)). The studies included in this review have identified spatial heterogeneity of WMHs with various impairments, diseases, and pathologies as well as with sex and (cerebro)vascular risk factors.

**Conclusions:** The results show that studying white matter hyperintensities on a more granular level might give a deeper understanding of the underlying neuropathology and their effects. This motivates further studies examining the spatial patterns of white matter hyperintensities.

## Introduction

White Matter Hyperintensities (WMHs) of presumed vascular origin are a widely studied marker of cerebral small vessel disease (SVD).^1^ This disease burden appears hyperintense on T2-weighted Magnetic Resonance Imaging (MRI), and is often characterized on FLuid Attenuated Inversion Recovery (FLAIR) imaging.^1^ Studies have demonstrated that their prevalence and severity increase with age.^2,3^ Moreover, it has been demonstrated that this cerebrovascular disease burden is associated with various impairments, diseases, and pathologies, such as motor^4^ and mood disorders,^5,6^ cognitive impairment (CI),^7–9^ dementia (DEM),^10,11^ and stroke.^12,13^ Additionally, the presentation of WMHs in the brain are associated with sex^14^ and clinical factors including (cerebro)vascular risk factors like hypertension (HTN)^15^ and diabetes type 2 (DM2).^16^ Due to its high prevalence, multiple studies have reviewed the evidence on its prevalence and modifying factors over the years.^8,17–21^

WMH burden is heterogeneous in location and size and appears as punctate, focal, and/or confluent lesions.^22^ It is commonly characterized using semi-quantitative visual rating scales, such as Fazekas,^22^ Manolio,^23^ or Scheltens,^24^ or by using fully-quantitative volumetric measurements based on either manual, semi-automated, or fully-automated approaches. To date, however, there is no universally established methodology for quantification, as their utility depends on availability, time costs, and quality of the imaging data.^17^ However, with the increased prevalence of fully automated and/or deep learning-enabled methodology, volumetric evaluations in increasingly larger cohorts are becoming more prevalent.^14,17,25,26^

While most investigations tend to summarize WMH burden as a single volumetric measure, researchers have started to acknowledge the importance of its spatial distributions to gain additional insights into the underlying neuropathology.^27^ Several studies have therefore examined the spatial patterns of WMHs in various populations. The aim of this systematic review is to give an overview of the evidence demonstrating pathological, clinical, and cerebrovascular risk factor effects on spatial patterns of WMHs in adult populations and clinical cohorts, by summarizing the increasing evidence of spatial specificity with respect to WMH burden.

## Methods

This systematic review was performed using the Preferred Reporting Items for Systematic Reviews and Meta-Analysis (PRISMA) Statement.^28^ This review was not registered and no review protocol was prepared. The associated PRISMA checklist can be found in the appendix.

### Search Strategy and Study Selection Criteria

Studies have been identified by an advanced search on PubMed. The in STRIVE^1^ described naming conventions for WMHs were utilized as a reference to include the most prominent terms for WMHs. Additionally, we restricted our analysis to studies with MRI FLAIR data published before February 1st, 2023. Due to the lack of consensus of nomenclature for the investigation of spatial WMH burden features, the terms “pattern”, “topology”, “topography” and “spatial” were included. The full search string is given as: (White Matter Hyperintensity OR White Matter Lesion OR White Matter Disease OR Leukoaraiosis) AND (MRI AND (FLAIR OR (Fluid Attenuated Inversion Recovery))) AND (spatial OR pattern OR topology OR topography). Only articles written in English were considered.

All abstracts were subsequently screened for eligibility. Studies were limited to human adults (>18 years) with sample sizes greater than 20 participants/patients that investigated whole-brain WMH patterns and their relation to risk factors and diseases. Studies investigating multiple sclerosis or tuberous sclerosis were not considered, following the STRIVE recommendation.^1^ Descriptive studies without the aim to describe the association of the observed patterns were excluded for the purpose of this review. Finally, we extended our selection by examining the cited literature of the identified studies for additional relevant articles.

### Data collection

The screening was performed by one reviewer without the use of automated tools (J.B.). The final decision over study inclusion was reached in consensus with a second reviewer (M.D.S.). Data were extracted using a standardized form that captured (1) disease type(s), (2) the number of patients of each disease type, (3) age, (4) sex, (5) the quantification method, and (6) spatial pattern analysis (see Table 1).

**Table 1:**
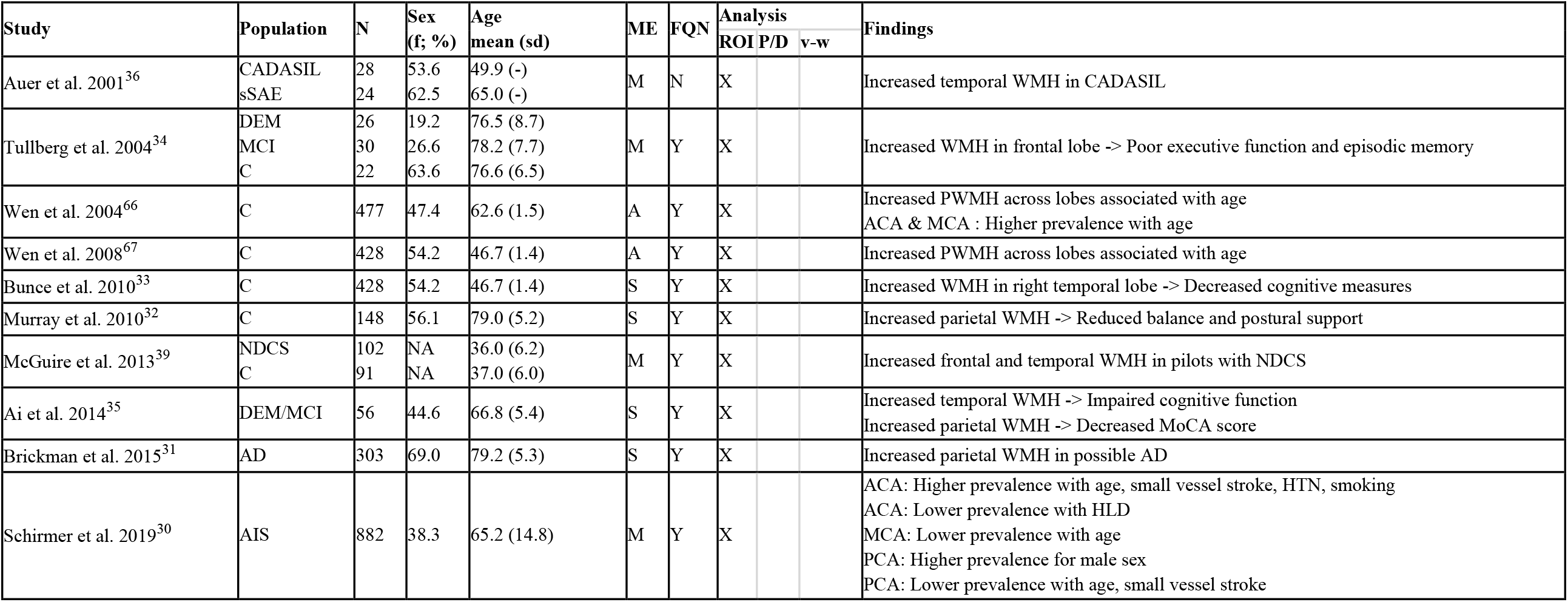
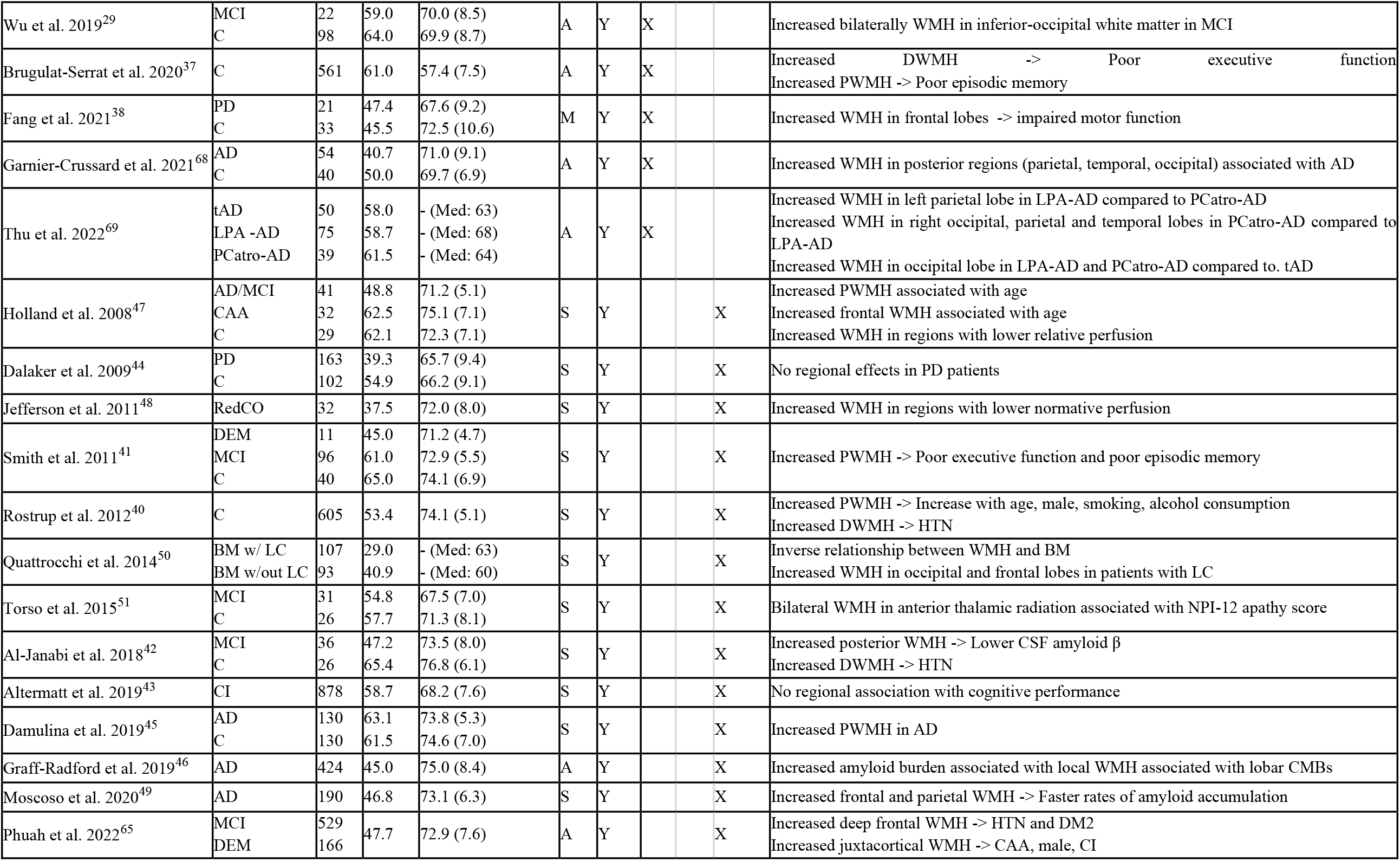
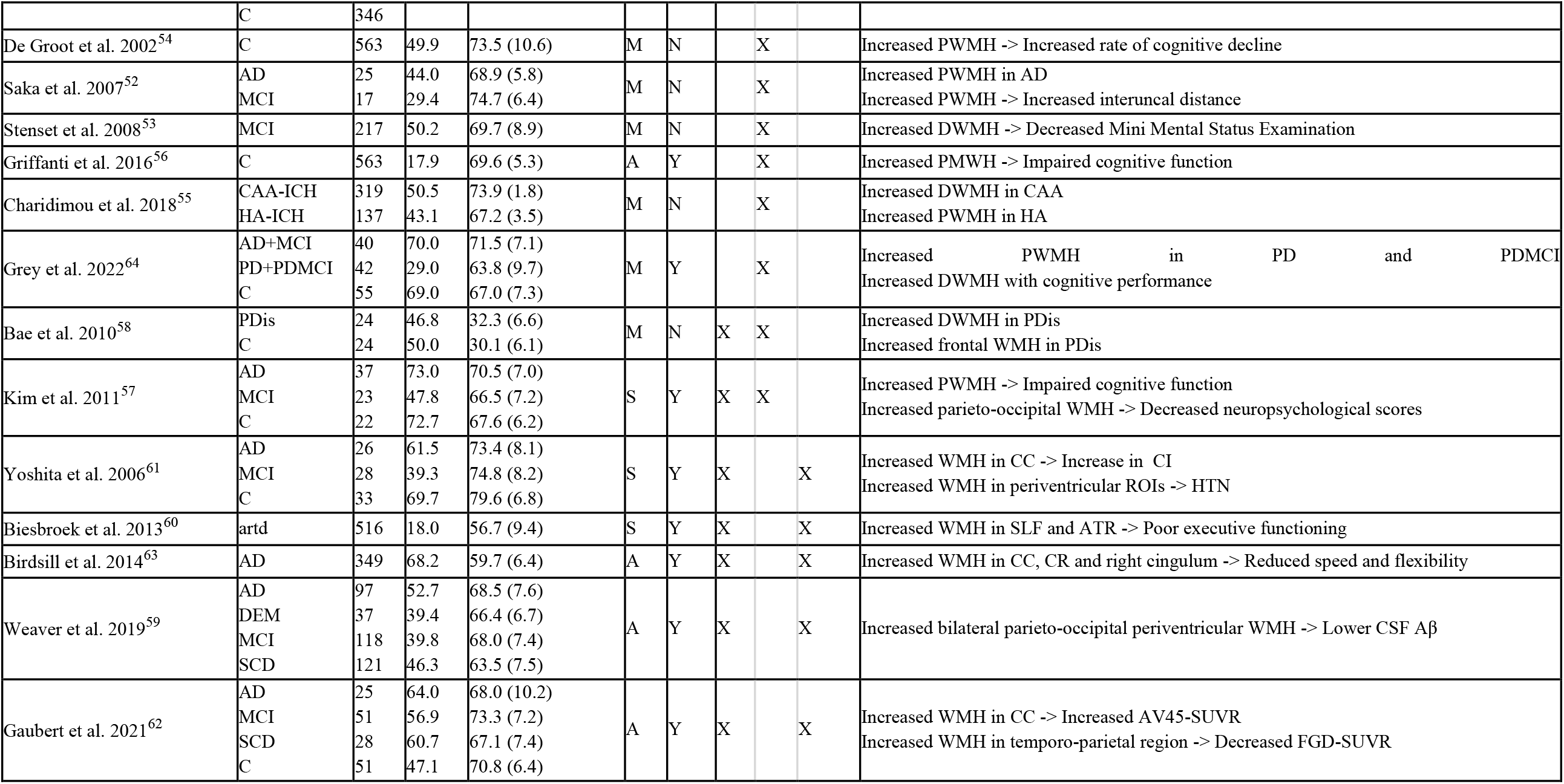
Overview of the study cohorts investigating spatial WMH burden patterns and summary of their findings. A = automatic, ACA = Anterior Cerebral Artery, AD = Alzheimer’s Disease, AIS = Acute Ischemic Stroke, artd = arterial disease, ATR = Anterior, Thalamic Radiation, AV45 = Florbetapir, BM = Brain Metastases, C = Control, CAA = Cerebral Amyloid Angiopathy, CADASIL = Cerebral Autosomal Dominant Arteriopathy with Subcortical Infarcts and Leukoencephalopathy, CC = Corpus Callosum, CI = Cognitive Impairment, CMBs = Cerebral Microbleeds, CR = Corona Radiata, CSF = Cerebrospinal Fluid, DEM = Dementia, DM2 = Diabetes Mellitus type 2, f = female, FDG = Fluordesoxyglucose, FQN = Fully Quantitative, HA = Hypertensive Arteriopathy, HLD = Hyperlipidemia, HTN = Hypertension, ICH = Intracerebral Hemorrhage, LC = Lung Cancer, LPA-AD =logopenic progressive aphasia, M = manual, MCA = Middle Cerebral Artery, MCI = Mild Cognitive Impairment, ME = Method, MoCA = Montreal Cognitive Assessment, N = cohort size, NDCS = Neurologic Decompression Sickness, NPI = Neuropsychiatric Inventory, P/D = PWMH/DWMH, PCA = Posterior Cerebral Artery, PCatro-AD = posterior cortical atrophy, PD = Parkinson Disease, PDis = Panic Disorder, RedCO = Reduced Cardiac Output, ROI = Region of Interest, S = semi-automatic, SCD = Subjective Cognitive Decline, sd = standard deviation, SLF = Superior Longitudinal Fasciculus, sSAE = sporadic Subcortical Arteriosclerotic Encephalopathy, SUVR = Standardised Uptake Value Ratio, tAD = typical amnestic Alzheimer’s Disease, v-w = voxel-wise

## Results

A total of 380 studies were identified by the initial literature search, of which 41 studies satisfied the inclusion criteria (see Figure 1 for a detailed description of the selection phase).^29–69^ These studies included cohorts based on mild cognitive impairment (MCI) (15/41), Alzheimer’s Disease (AD) (14/41), Dementia (DEM) (5/41), Parkinson’s Disease (PD) (3/41), and subjective cognitive decline (SCD) (2/41). Six of the 41 studies investigated cognitively normal cohorts, two of which were population-based.^32,37^ Additionally one study described the spatial WMH patterns in an acute ischemic stroke (AIS) cohort^30^ and one in a population with reduced cardiac output (RedCO)^48^. Cohorts ranged from 32 to 882 patients/participants (median cohort size: 191.5, 51.6% female (range 17.9 - 81.3 %)). The mean age of the investigated cohorts ranged from 30 to 80 years (median: 69.9 years; mean age not reported in two studies^50,69^).

**Figure 1:**
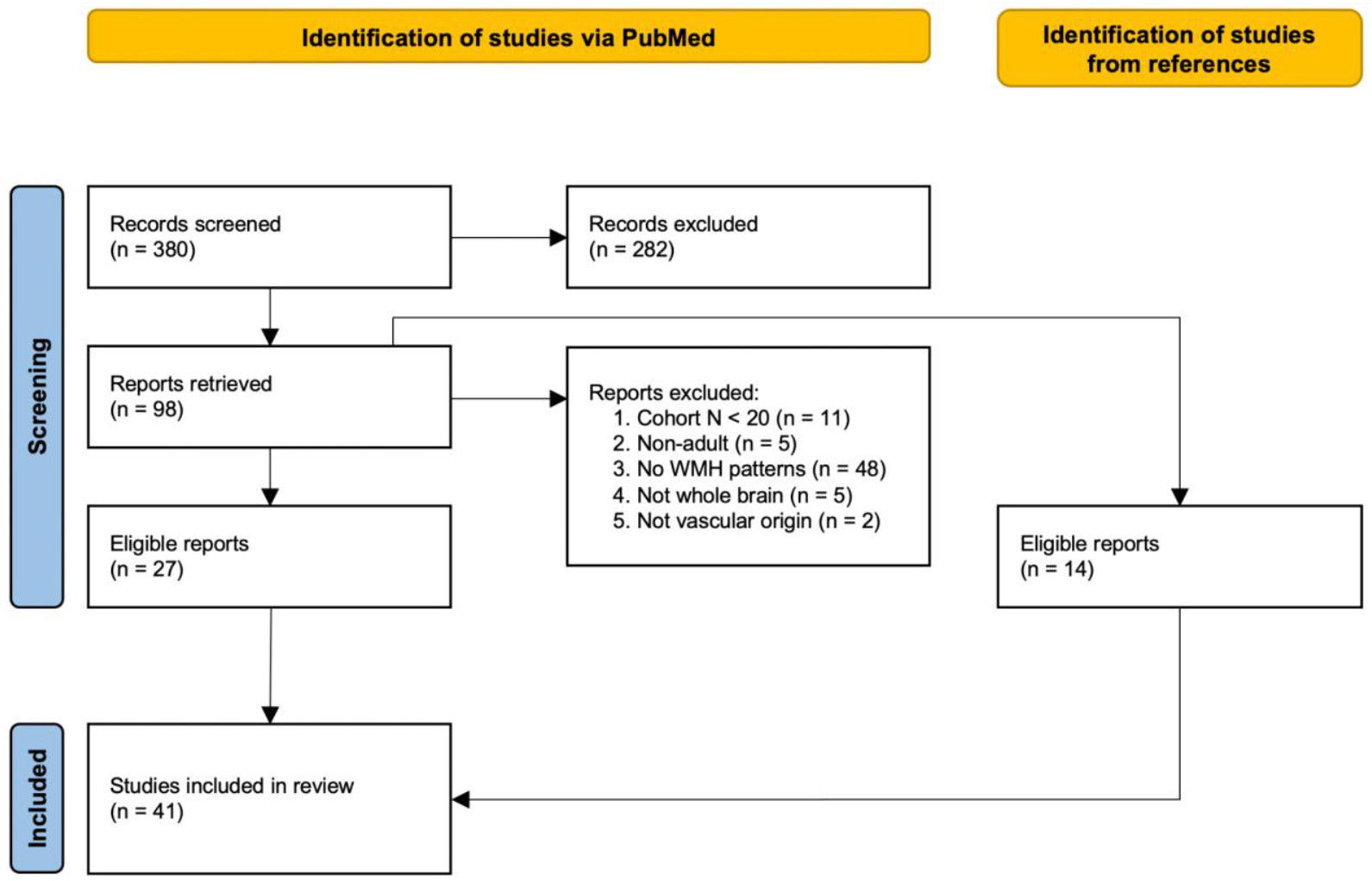
Flow-chart of study selection based on the PRISMA statement.^28^ Studies were identified through an advanced search on PubMed. First, abstracts were screened, followed by reading the retrieved publications and excluding non-relevant studies. Exclusion criteria - Reason 1: Cohort included less than 20 subjects. Reason 2: Study was not performed in adult populations (>18 years). Reason 3: Study did not investigate spatial patterns of WMHs (e.g., studies investigated spatial patterns of other markers, such as activation patterns of functional MRI, only included total WMH burden as a covariate, or fully descriptive studies). Reason 4: Study did not assess the whole brain for analysis. Reason 5: Study investigating multiple sclerosis^70^ or tuberous sclerosis^71^ which do not meet the STRIVE^1^ definition of WMH. The reference list of each retrieved report was additionally examined for eligible studies.

### WMH Quantification

To study spatial features of WMH burden and its association with clinical correlates and risk factors, it is necessary to characterize the extent of the burden. In the identified studies, WMHs were assessed by employing qualitative/semi-quantitative (n=6), or fully quantitative approaches (n=35). Most studies utilized either semi- or fully-automated WMH segmentation methodology (31/35), eleven of which were based on respective in-house developed algorithms, six using the Lesion Segmentation Toolbox (LST),^72^ and the remainder employed other available tools like the Brain Intensity AbNormality Classification Algorithm (BIANCA)^73^ or the Medical Image Processing, Analysis, and Visualization (MIPAV) software package.^74^ In five studies lesions were manually delineated.

#### Semi-quantitative/qualitative

Semi-quantitative/qualitative measures represent the degree of lesional dispersion and severeness with help of visual rating scales. These scales provide a way to measure WMH burden and describe its topology without explicit lesion delineation. The used scales were the Fazekas^22^ (and its derivatives; n=5), and Scheltens^24^ (n=1) scales. The latter incorporates WMH topology in the form of local, lobar burden, while the Fazekas scale is employed for whole-brain evaluation.

#### Fully-quantitative

Fully-quantitative approaches describe the disease burden as a volumetric measure, e.g., in cubic centimeters or milliliters. WMHs are delineated either manually, semi-, or fully-automatically. Manual WMH segmentation, while often considered the gold standard in the field, is time-consuming and shows high intra- and inter-rater variability.^75^ To address this challenge, many automated assessment algorithms have been developed,^25^ however, no universally applicable algorithm exists.^17^

### Spatial Pattern Analyses

Based on the identified literature, we distinguish between three analysis approaches for studying spatial patterns of WMH burden: 1) Region of interest (ROI), 2) periventricular (PWMH) and deep WMH (DWMH), and 3) voxel-wise analysis. A comprehensive overview of the included studies and their findings is given in Table 1.

#### Regions of interest

In this approach, the brain is subdivided into ROIs, based on brain areas that individual studies hypothesize to be associated with specific biomarkers. These ROIs may represent, e.g. cortical lobes,^29,31–33,35,36,38,39,66–69^ or other specific regions that may be relevant to the patient cohort or disease under investigation, e.g. using vascular territories.^30,66^ To identify and define ROIs, studies often rely on image registration to a brain template, which is often derived for the general population or a control cohort, and on which the ROIs are defined.

We identified eleven studies using ROI analyses.^29–39,66–69^ Of these studies eight utilized the definition of cortical lobes or a modified version of them as ROIs. While increased parietal WHM burden was associated with AD,^31^ reduced balance and postural support,^32^ and with poor cognitive function, measured with the Montreal cognitive assessment (MoCA),^35^ increased WMH burden in the frontal lobe was related to poor executive function and episodic memory^34^ in patients with PD^38^ and neurological decompression sickness (NDCS).^39^ Increased temporal WMH burden corresponded with decreased cognitive measures,^33^ and impaired cognitive function,^35^ while another study associated increased inferior-occipital WMH burden with MCI.^29^ Additionally, Garnier-Crussard et al.^68^ and Thu et al.^69^ found WMH in the posterior regions (parietal, temporal, and occipital) to be associated predominantly with AD. Thu et al.^69^ further compared the local WMH burden across atypical variants of AD logopenic progressive aphasia (LPA-AD), posterior cortical atrophy (PCatro-AD) and typical amnestic AD (tAD). Patients with LPA-AD compared to PCatro-AD showed a higher left/right posterior lobar WMH burden, and patients with either atypical variants had more WMHs in the occipital lobes compared to patients with tAD. In patients with cerebral autosomal dominant arteriopathy with subcortical infarcts and leukoencephalopathy (CADASIL), Auer et al.^36^ found an increased WMH burden in the temporal lobes, and in those with NDCS, McGuire et al.^39^ described an increased WMH burden in the frontal and temporal lobes. Schirmer et al.^30^ demonstrated a shift of predominance of WMH burden in a cohort of AIS patients and by using areas of cortical blood flow, i.e. vascular territories, that were associated with age, sex, small vessel stroke, hypertension, smoking, and hyperlipidemia. Similarly, Wen et al.^66^ introduced cortical blood flow regions in addition to lobar ROIs. They found increased WMH burden in the anterior and middle cerebral artery regions as well as in the periventricular regions. The association between advanced age and increased periventricular WMH burden was confirmed by a follow-up study^67^ of younger patients (mean age under 50 years). Finally, using a bullseye representation, Tullberg et al.^34^ showed an increased WMH burden in the deep white matter which was associated with poor executive function whereas increased WMH burden close to the ventricles was found to be associated with poor episodic memory. While this approach roughly relates to P/DWMH, discussed in the next section, they did not follow the common definitions for stratification in their work, resulting in 36 unique ROIs.

#### PWMH and DWMH

PWMH and DWMH are umbrella terms for multiple definitions in the literature, all relating WMH burden to the distance with respect to the ventricular surface.^56^ In general, PWMHs are adjacent or in close proximity to the lateral ventricles, while DWMHs are located at larger distances inside the subcortical white matter. While PWMH and DWMH can be considered ROIs, this definition aims to reflect different functional, histopathological, and etiological features of WMH burden.

We identified six studies distinguishing PWMH and DWMH.^52–56,64^ Increased PWMH burden was found to be associated with PD,^64^ AD and brain atrophy,^52^ faster progression in cognitive decline,^54^ and impaired cognitive function.^56^ Increased DWMH burden was associated with a decreased mini mental status examination score in patients with MCI.^53^ Another study showed increased DWMH burden in subjects with intracerebral hemorrhage and cerebral amyloid angiopathy (CAA), while subjects with intracerebral hemorrhage and hypertensive arteriopathy (HA) presented with increased PWMH burden.^55^

#### Voxel-Wise

In the voxel-wise approach, each white matter voxel is studied for the presence of WMHs. After image registration to a template, the voxel-based frequency can be used to either derive maps showing clusters of WMHs within a specific cohort or serve as the basis for voxel-based lesion-symptom mapping. The advantage of voxel-wise analysis lies in the hypothesis-free, data-driven localization of lesion clusters compared to ROI approaches. Furthermore, it enables studies to combine this analysis with studying associations with other localized cerebrovascular information, such as presence of cerebral microbleeds and perfusion.

Thirteen studies utilized a voxel-wise analysis approach.^40–51,65^ In patients with AD and MCI, local WMH burden in the frontal and parietal lobes close to the ventricles was associated with an increased amyloid burden.^42,46,49,65^ Increased WMH burden in voxels around the ventricles was found to be associated with advanced age,^40^ poor executive function and episodic memory,^41^ and with AD.^45^ Further, juxtacortical WMH load was found to be associated with CI, male sex, and CAA, and deep frontal WMH burden with hypertension and DM2.^65^ Quattrocchi et al.^50^ showed that patients with lung cancer (LC) with brain metastases (BM) presented with increased WMH burden in the frontal and occipital lobes, compared to those in a control group. They also identified an inverse relationship between the presence of BM and WMH burden. Two additional studies measured local perfusion and demonstrated an inverse relationship between WMH burden and perfusion, meaning higher WMH burden related to low relative perfusion in the normal appearing white matter.^47,48^ In another study increased WMH burden bilaterally in the anterior thalamic radiation was found to be associated with apathy.^51^ Lastly, two studies reported no regional associations between WMH burden and cognitive performance^43^ or PD.^44^

#### Mixed Approach

Seven studies utilized a mixed spatial analysis approach, two with ROI as well as distinguishing between PWMH and DWMH,^57,58^ and five studies applying ROI and voxel-wise analyses.^59–63^ Kim et al.^57^ found increased PWMH burden associated with impaired cognitive function and increased parieto-occipital WMH burden associated with decreased neuropsychological scores. Bea et al.^57^ found increased frontal and deep WMH burden in patients with panic disorder compared to controls. Four of the studies using ROI and voxel-wise analyses found local associations between WMH burden and lower cerebrospinal fluid (CSF) amyloid burden,^59^ poor executive function,^60^ increased vascular risk factors and CI,^61^ and reduced speed and flexibility of executive function.^63^ Gaubert et al.^62^ demonstrated associations of regional WMH distributions in multiple locations, such as the posterior lobes and the corpus callosum, with multimodal brain biomarkers of AD.

#### Spatial effects of vascular risk factors and clinical correlates

Some of the discussed studies reported spatial effects of vascular risk factors and clinical correlates, such as age, sex, HTN, smoking, alcohol, stroke, hyperlipidemia (HLD), and specific neurodegenerative and other diseases.

Advanced age was associated with increased PWMH^47,53,56,66,67^ and DWMH^29,53^ burden as well as with increased WMH burden in the parietal lobes.^47^ Furthermore - using vascular territories as ROIs - an association was found between advanced age and increased and decreased relative WMH burden in the anterior cerebral artery (ACA) and middle cerebral artery (MCA) territory, respectively.^30,66^ Men showed increased PWMH^40^ and WMH burden in the posterior cerebral artery (PCA) region,^30^ while women presented with increased frontal WMH burden.^33^ Increased DWMH^40,42,61^ and ACA territory burden^30^ were associated with HTN. Smoking^30^ and alcohol consumption^40^ showed increased PWMH burden, while smoking also showed increased WMH burden in the ACA territory.^30^ Patients with small vessel stroke presented with increased WMH burden in the ACA and decreased WMH burden in the PCA territory, while HLD led to a decreased WMH burden in the ACA territory.^30^ Patients with AD showed a broad spectrum of spatial effects: they had increased WMH burden in the frontal^34,46,49^ and parietal^31,34,46,49^ lobes, in the posterior cerebrum,^62,68,69^ the corpus callosum,^61–63^ the corona radiata,^62,63^ the anterior thalamic radiation^62,63^ as well as increased PWMH^45,46,52,57,59,61^ and DWMH^53^ burden. Patients with MCI showed similar distributions with increased WMH burden in the parietal lobe,^35^ the posterior cerebrum^42^ and increased PWMH burden,^41,61^ but also increased WMH burden in the temporal lobe,^35^ which was also found to be associated with CADASIL.^36^ Patients with PD presented with increased WMH burden in the frontal lobe.^38^ Other diseases like arterial disease showed increased WMH burden in the superior longitudinal fasciculus and the anterior thalamic radiation,^60^ panic disorder was associated with increased DWMH burden as well as with increased WMH burden in the frontal lobe.^58^ NDCS^39^ and LC with BM^50^ were associated with WMH burden in the frontal lobe. CAA and HA were associated with increased DWMH and PWMH burden, respectively.^55^ The associations are summarized in Table 2.

**Table 2.** Vascular risk factors and clinical correlates affecting spatial patterns. Abbreviations: ACA = Anterior Cerebral Artery, AD = Alzheimer’s Disease, ATR = Anterior Thalamic Radiation, artd = arterial disease, BM = Brain Metastases, CAA = Cerebral Amyloid Angiopathy, CADASIL = Cerebral Autosomal Dominant Arteriopathy with Subcortical Infarcts and Leukoencephalopathy, CC = Corpus Callosum, CR = Corona Radiata, HA = Hypertensive Arteriopathy, HLD = Hyperlipidemia, HTN = Hypertension, LC = Lung Cancer, MCA = Middle Cerebral Artery, MCI = Mild Cognitive Impairment, NDCS = Neurologic Decompression Sickness, PCA = Posterior Cerebral Artery, PD = Parkinson’s Disease, PDis = Panic Disorder, SLF = Superior Longitudinal Fasciculus

| Vascular risk factors and clinical correlates | Spatial effect [# Studies] |
| --- | --- |
| Age | Increased PWMH [n=5]<sup>47,53,56,66,67</sup><br>Increased DWMH [n=2]<sup>29,53</sup><br>Increased parietal WMH [n=1]<sup>47</sup><br>Increased WMH in ACA [n=2]<sup>30,66</sup><br>Decreased WMH in MCA [n=2]<sup>30,66</sup> |
| Sex | Increased PWMH (male<sup>40,65</sup>) [n=2]<br>Increased frontal WMH (female<sup>33</sup>) [n=1]<br>Increased WMH in PCA (male<sup>30</sup>) [n=1] |
| Vascular risk Factors | Increased DWMH (HTN<sup>40,42,61</sup>) [n=3]<br>Increased PWMH (smoking<sup>30</sup>, alcohol<sup>40</sup>) [n=2]<br>Increased relative WMH in ACA (HTN, smoking, stroke)<sup>30</sup> [n=1]<br>Decreased relative WMH in ACA (HLD<sup>30</sup>) [n=1]<br>Decreased relative WMH in PCA (stroke<sup>30</sup>) [n=1] |
| Neurodegenerative diseases | Increased frontal WMH (AD<sup>34,46,49</sup>, PD<sup>38</sup>) [n=4]<br>Increased temporal WMH (MCI<sup>35</sup>, CADASIL<sup>36</sup>) [n=2]<br>Increased parietal WMH (AD<sup>31,34,46,49</sup>, MCI<sup>35</sup>) [n=5]<br>Increased posterior WMH (AD<sup>62,68,69</sup>, MCI<sup>42</sup>) [n=4]<br>Increased WMH in CC (AD<sup>61-63</sup>) [n=3]<br>Increased WMH in CR (AD<sup>62,63</sup>) [n=2]<br>Increased PWMH (AD<sup>45,46,52,57,59,61</sup>, MCI<sup>41,61</sup>, PD<sup>64</sup>) [n=9]<br>Increased DWMH (AD<sup>53</sup>, MCI<sup>29</sup>) [n=2]<br>Increased WMH in ATR (AD<sup>51</sup>) [n=1] |
| Other diseases | Increased DWMH (PDis<sup>58</sup>, CAA<sup>55</sup>) [n=2]<br>Increased PWMH (HA<sup>55</sup>) [n=1]<br>Increased juxtacortical WMH (CAA<sup>65</sup>) [n=1]<br>Increased frontal WMH (PDis<sup>58</sup>, NDCS<sup>39</sup>, DM2<sup>65</sup>, LC&BM<sup>50</sup>) [n=4]<br>Increased WMH in SLF (artd<sup>60</sup>) [n=1]<br>Increased WMH in ATR (artd<sup>60</sup>) [n=1] |

## Discussion

WMH burden has been linked to various impairments, diseases, pathologies, as well as (cerebro)vascular risk and clinical factors. Subsequently, it has been used as a biomarker in multiple studies, often summarized as total WMH burden or load.^8^ However, WMHs demonstrate spatial distributions in the brain that are specific to diseases and other clinical and risk factors. Investigation of the topographical aspects of this marker of cerebral small vessel disease, rather than summarizing it as a single, volumetric measure, can therefore give new insights into the underlying pathophysiology and studied correlates of interest.

This systematic review of 41 studies gives new insights into the topography of WMHs as a biomarker and provides strong evidence for the importance and benefit of studying their spatial distribution. In general, we differentiate between qualitative and semi-/quantitative WMH characterization approaches. While qualitative approaches are easy to perform manually in smaller cohorts, other approaches, such as probabilistic segmentation methods, can be utilized to extend the analyses to bigger cohort sizes. Importantly, with the event of deep learning enabled pipelines for white matter hyperintensity segmentation, fully automated, quantitative characterization has become more prominent in the literature. The latter has the benefit of removing inter-rate variability within a study, however, comparability of segmentation between studies may not be given, if different algorithms are used, due to the high heterogeneity of performance.^25^

For spatial stratification, we identified three general approaches to studying the spatial patterns of WMH. These included ROI, PWMH/DWMH, and voxel-wise stratification. While a generally accepted methodology on how to categorize WMH is lacking, the described relationships with risk factors and clinical correlates demonstrated consistent trends. In general, it is difficult to define such a gold-standard for spatial analyses, as both hypothesis (ROI) and hypothesis-free (voxel-wise) approaches have merit and depend on the specific research question that is being answered. An ROI approach may yield new insights and potentially simplify the analysis, however, the selection of ROI definition needs to be based on the underlying biological hypothesis. A voxel-wise approach provides a more granular topographical investigation through a data driven approach, however, the resulting interpretation of the results should consider the anatomical structure of the brain, and potentially relate its findings back to commonly used ROIs to facilitate the interpretation of the findings. Despite the different approaches and use of varying WMH quantification techniques, the discussed findings presented in this review agreed in the direction of the found association, suggesting the presence of a biologic signal.

We defined WMH in this review according to the STRIVE criteria^1^, i.e. hyperintense signal in the white matter of presumed vascular origin, assessed using FLAIR imaging. However, this systematic review covered populations with various diseases, including AD, DEM and PD, for which, at least in part, it is still unclear whether SVD is etiologically involved in the neurodegenerative processes or occurs coincidentally. For example, it has been suggested that SVD might precede Parkinsonism related pathology,^76^ whereas WMH and amyloid accumulations might be independent, but additive processes.^77^ Nevertheless, it is established that WMH are highly prevalent in these neurodegenerative diseases and play a key role in cognitive impairment, warranting their investigation.^7–9^

The studies included in this systematic review investigated spatial patterns of WMH in populations with different diseases. Spatial stratification was predominantly performed in studies investigating cognitive impairment (n=23), identifying differences in spatial patterns of WMH in PWMH (n=7; for AD and MCI),^41,45,46,52,57,59,61^ as well as in the parietal (n=5; for AD and MCI),^31,34,35,46,49^ posterior (n=4; for AD and MCI),^42,62,68,69^ and frontal (n=3; for AD)^34,46,49^ lobar regions. In this particular study population, spatial patterns of WMH appear across the entire brain. However, given the high overall global disease burden in these patient groups, as well as heterogeneity of confounding factors that are present and methodological approaches for characterizing WMH burden used, it is difficult to identify a consensus on distinct, disease specific spatial patterns.

Three studies identified distinctive spatial WMH patterns for different subtypes of SVD.^36,55,65^ While juxtracortical WMH were associated with probable CAA,^55^ deep (frontal) WMH were predominantly found in patients with risk factors for arteriosclerosis,^65^ and temporal WMH with CADASIL.^36^ Again, the methodological approaches differed across studies, hindering a direct comparison. Another three studies explored spatial patterns of WMH in patients with PD.^38,44,64^ While one of them did not identify spatial patterns specific to patients with PD,^44^ Fang et al.^38^ found an increased WMH burden in the frontal lobe, and Grey et al.^64^ identified increased PWMH burden. The differences in these findings may be explained due to the variations in age and sex distribution of the study populations, as well as the different methodology used for characterizing WMH. Additional studies are needed for these and other common diseases, such as stroke, to fully explore the topography of the cerebrovascular disease burden. Future research may also consider elucidating differences, not only between patient cohorts compared to the general population, but between patient cohorts of various diseases to further highlight the applicability of spatially specific disease burden as biomarkers for early disease prognostication.

Epidemiologic research over the past decades has highlighted the importance of vascular risk factors in WMH.^3,19,75^ Overall, studies have identified spatial heterogeneity of WMHs related to risk factors and clinical correlates. Localized WMH burden was found to be positively correlated with vascular risk factors, such as HTN, alcohol consumption, and history of smoking, as well as clinical correlates, such as sex (see Table 2). The studies described in this review also demonstrated that patients with HTN mainly presented with an increase in DWMH burden, whereas smoking and alcohol consumption were linked to an increase in PWMH. This supports the underlying assumption that deep and periventricular WMH are different etiologies that are individually affected by different vascular risk factors.

While only a few studies specifically investigated sex differences, the research findings highlighted in this systematic review suggest that it plays an important role in the spatial distribution of WMH, with men showing higher WMH in periventricular and PCA regions and women with higher frontal burden. Sex differences in total WMH burden, as well as differences in women pre- and post-menopausal, have been shown previously in the literature.^3,14,78–80^ These findings, with the above described spatial differences in disease burden, however, have not been fully explored, but should be accounted for in future studies.

Our systematic review further identified different spatial patterns of WMHs associated with poor overall cognitive outcomes. The results, however, are inconsistent and warrant further, systematic studies. Only a few studies (n=3) investigated different cognitive domains, specifically looking into spatial patterns in relation to executive function and episodic memory. While there were no distinctive WMH patterns in studies of older participants who ranged from cognitively normal to demented, a study in middle-aged, cognitively normal participants found a distinct disease burden topography. Specifically, Brugulat-Serrat et al.^37^ showed that cognitively unimpaired, middle-aged adults with an increase in DWMH burden performed poorer in tasks relating to executive memory, whereas increased PWMH burden was related to poor episodic memory. In other studies in older adults, Tullberg et al.^34^ demonstrated that poor executive function and episodic memory was related to increased frontal WMH, whereas Smith et al.^41^ found that these were associated with increased PWMH. While these findings are not exclusive of one another, however, the use of different definitions for spatial stratification hinders a direct comparison. Both studies suggest, however, that this spatial distinction might be an early marker for cognitive impairment in younger people, where the overall WMH burden in the brain is still relatively low. Nonetheless, further studies across the lifespan are needed to fully examine the described association.

While research into spatial patterns of WMH is a rapidly developing field that first emerged around 20 years ago with most publications in the last decade, a commonly agreed terminology does not yet exist, which hinders the identification of related literature. While our research terms included a large variety of descriptive words for WMH and topology, it is possible that studies were missed due to the large variety of nomenclature. Efforts reflecting other standardization approaches, such as STRIVE,^1^ may be warranted to significantly enhance the development of the field. In this systematic review, the varying definitions for spatial WMH burden did not allow to pool sufficient data for additional meta-analysis. Future research and meta-analyses would also significantly benefit from a consensus for standard terminology and of image analysis methodology for spatial patterns analyses of WMH.

In this systematic review, we discussed the evidence in the selected literature for spatial WMH patterns. While the data of the included studies did not allow for a quantitative synthesis and risk assessment of bias of results, intrinsic biases of the discussed studies exist. Investigating spatially stratified disease burden necessitates large cohort sizes to account for the general heterogeneity of WMH presentation. Here, cohort sizes varied significantly, ranging from 32 to 882 patients/participants, with most studies having cohort sizes of N<200 (median cohort size 191.5). While smaller cohorts may be sufficient to provide first insights into spatial patterns of WMH, larger cohort sizes are likely necessary to fully uncover the driving factors in observed WMH topography, its associated risk factors and clinical correlates. Moreover, studies significantly varied in populations under investigation, with a total of 16 different diagnoses, in addition to “control” groups, with a range of 1 - 15 studies per diagnosis. Studies including MCI patients were most prevalent. As described above, more disease specific evidence is required to remove uncertainties in the results that are present due to intrinsic biases in the study cohorts. Additionally, most studies included in this systematic review were conducted in older populations (median cohort age: 69.9 years), specifically unraveling spatial patterns of WMH with respect to AD, MCI and normal cognitive function. In general, WMH is most prominent in aging populations, as age is a key determinant of overall WMH burden, enabling easier automated quantification, where small errors in segmentation accuracy often only relate to minute variations in relative burden. Only a few studies investigated spatial patterns of WMH in younger participants, however, these investigations focused on rather specific study populations, e.g. presenting with panic disorders or pilots. The results presented here may therefore not generalize well to the general population. Studies are warranted to disentangle spatial effects of WMH in younger people and populations with lower WMH burden from the general population, as well as overall population studies which include a wide age range.

This systematic review provides an overview of the state of spatial patterns of WMHs across cohorts in adults. While in this review, we only extracted literature from a single database (PubMed), it provides a good starting point for the emerging studies. Here, we focused on adults, but WMH can emerge earlier in life and is often linked with disease state. Overall this systematic review has met its expectations and achieved its objectives.

## Data Availability

All data produced are available online at

## Acknowledgements

This project has received funding from the BONFOR program of the Medical Faculty of the Friedrich-Wilhelms University Bonn (O-194.0001). VL is supported by the Marga and Walter Boll Foundation, Kerpen, Germany.

## Author contributions

All authors had full access to all the data in the study and take responsibility for the integrity of the data and the accuracy of the data analysis. Conceptualization, JB, VL, MS; Methodology, JB, VL, MS; Investigation, JB, MS; Formal Analysis, JB; Writing – Original Draft, JB, VL, MS; Writing – Review & Editing, JB, VL, MS; Visualization, JB; Supervision, MS;

## References

1. Wardlaw JM, Smith EE, Biessels GJ, et al. Neuroimaging standards for research into small vessel disease and its contribution to ageing and neurodegeneration. Lancet Neurol. 2013;12(8):822–838. doi:10.1016/S1474-4422(13)70124-8

2. Morris Z, Whiteley WN, Longstreth WT, et al. Incidental findings on brain magnetic resonance imaging: systematic review and meta-analysis. BMJ. 2009;339(aug17 1):b3016–b3016. doi:10.1136/bmj.b3016

3. Das AS, Regenhardt RW, Vernooij MW, Blacker D, Charidimou A, Viswanathan A. Asymptomatic Cerebral Small Vessel Disease: Insights from Population-Based Studies. J Stroke. 2019;21(2):121–138. doi:10.5853/jos.2018.03608

4. Smith EE, O’Donnell M, Dagenais G, et al. Early cerebral small vessel disease and brain volume, cognition, and gait. Ann Neurol. 2015;77(2):251–261. doi:10.1002/ana.24320

5. Lohner V, Brookes RL, Hollocks MJ, Morris RG, Markus HS. Apathy, but not depression, is associated with executive dysfunction in cerebral small vessel disease. PLoS ONE. 2017;12(5):e0176943. doi:10.1371/journal.pone.0176943

6. van Uden IWM, Tuladhar AM, de Laat KF, et al. White matter integrity and depressive symptoms in cerebral small vessel disease: The RUN DMC study. Am J Geriatr Psychiatry Off J Am Assoc Geriatr Psychiatry. 2015;23(5):525–535. doi:10.1016/j.jagp.2014.07.002

7. Croall ID, Lohner V, Moynihan B, et al. Using DTI to assess white matter microstructure in cerebral small vessel disease (SVD) in multicentre studies. Clin Sci Lond Engl 1979. 2017;131(12):1361–1373. doi:10.1042/CS20170146

8. Debette S, Markus HS. The clinical importance of white matter hyperintensities on brain magnetic resonance imaging: systematic review and meta-analysis. BMJ. 2010;341:c3666. doi:10.1136/bmj.c3666

9. METACOHORTS Consortium. Electronic address: joanna.wardlawed.ac.uk, METACOHORTS Consortium. METACOHORTS for the study of vascular disease and its contribution to cognitive decline and neurodegeneration: An initiative of the Joint Programme for Neurodegenerative Disease Research. Alzheimers Dement J Alzheimers Assoc. 2016;12(12):1235–1249. doi:10.1016/j.jalz.2016.06.004

10. Debette S, Beiser A, DeCarli C, et al. Association of MRI Markers of Vascular Brain Injury With Incident Stroke, Mild Cognitive Impairment, Dementia, and Mortality: The Framingham Offspring Study. Stroke. 2010;41(4):600–606. doi:10.1161/STROKEAHA.109.570044

11. Prins ND, van Dijk EJ, den Heijer T, et al. Cerebral White Matter Lesions and the Risk of Dementia. Arch Neurol. 2004;61(10):1531. doi:10.1001/archneur.61.10.1531

12. Gerdes VEA, Kwa VIH, ten Cate H, Brandjes DPM, Büller HR, Stam J. Cerebral white matter lesions predict both ischemic strokes and myocardial infarctions in patients with established atherosclerotic disease. Atherosclerosis. 2006;186(1):166–172. doi:10.1016/j.atherosclerosis.2005.07.008

13. Giese AK, Schirmer MD, Dalca AV, et al. White matter hyperintensity burden in acute stroke patients differs by ischemic stroke subtype. Neurology. 2020;95(1):e79–e88. doi:10.1212/WNL.0000000000009728

14. Lohner V, Pehlivan G, Sanroma G, et al. The Relation Between Sex, Menopause, and White Matter Hyperintensities: The Rhineland Study. Neurology. Published online June 29, 2022:10.1212/WNL.0000000000200782. doi:10.1212/WNL.0000000000200782

15. Longstreth WT, Manolio TA, Arnold A, et al. Clinical Correlates of White Matter Findings on Cranial Magnetic Resonance Imaging of 3301 Elderly People: The Cardiovascular Health Study. Stroke. 1996;27(8):1274–1282. doi:10.1161/01.STR.27.8.1274

16. Ferguson SC, Blane A, Perros P, et al. Cognitive Ability and Brain Structure in Type 1 Diabetes: Relation to Microangiopathy and Preceding Severe Hypoglycemia. Diabetes. 2003;52(1):149–156. doi:10.2337/diabetes.52.1.149

17. Caligiuri ME, Perrotta P, Augimeri A, Rocca F, Quattrone A, Cherubini A. Automatic Detection of White Matter Hyperintensities in Healthy Aging and Pathology Using Magnetic Resonance Imaging: A Review. Neuroinformatics. 2015;13(3):261–276. doi:10.1007/s12021-015-9260-y

18. Frey BM, Petersen M, Mayer C, Schulz M, Cheng B, Thomalla G. Characterization of White Matter Hyperintensities in Large-Scale MRI-Studies. Front Neurol. 2019;10:238. doi:10.3389/fneur.2019.00238

19. Wardlaw JM, Valdés Hernández MC, Muñoz-Maniega S. What are White Matter Hyperintensities Made of? J Am Heart Assoc Cardiovasc Cerebrovasc Dis. 2015;4(6):e001140. doi:10.1161/JAHA.114.001140

20. Herrmann LL, Le Masurier M, Ebmeier KP. White matter hyperintensities in late life depression: a systematic review. J Neurol Neurosurg Psychiatry. 2007;79(6):619–624. doi:10.1136/jnnp.2007.124651

21. Melazzini L, Vitali P, Olivieri E, et al. White Matter Hyperintensities Quantification in Healthy Adults: A Systematic Review and Meta-Analysis. J Magn Reson Imaging. 2021;53(6):1732–1743. doi:10.1002/jmri.27479

22. Fazekas F, Chawluk J, Alavi A, Hurtig H, Zimmerman R. MR signal abnormalities at 1.5 T in Alzheimer’s dementia and normal aging. Am J Roentgenol. 1987;149(2):351–356. doi:10.2214/ajr.149.2.351

23. Manolio TA, Kronmal RA, Burke GL, et al. Magnetic resonance abnormalities and cardiovascular disease in older adults. The Cardiovascular Health Study. Stroke. 1994;25(2):318–327. doi:10.1161/01.str.25.2.318

24. Scheltens P, Barkhof F, Leys D, et al. A semiquantative rating scale for the assessment of signal hyperintensities on magnetic resonance imaging. J Neurol Sci. 1993;114(1):7–12. doi:10.1016/0022-510x(93)90041-v

25. Kuijf HJ, Biesbroek JM, De Bresser J, et al. Standardized Assessment of Automatic Segmentation of White Matter Hyperintensities and Results of the WMH Segmentation Challenge. IEEE Trans Med Imaging. 2019;38(11):2556–2568. doi:10.1109/TMI.2019.2905770

26. Schirmer MD, Dalca AV, Sridharan R, et al. White matter hyperintensity quantification in large-scale clinical acute ischemic stroke cohorts - The MRI-GENIE study. NeuroImage Clin. 2019;23:101884. doi:10.1016/j.nicl.2019.101884

27. Veluw SJ van, Barkhof F, Schirmer MD. White Matter Hyperintensity Spatial Patterns Provide Clues About Underlying Disease: Location Matters! Neurology. 2022;99(23):1017–1018. doi:10.1212/WNL.0000000000201398

28. Page MJ, McKenzie JE, Bossuyt PM, et al. The PRISMA 2020 statement: an updated guideline for reporting systematic reviews. BMJ. 2021;372:71. doi:10.1136/bmj.n71

29. Wu D, Albert M, Soldan A, et al. Multi-atlas based detection and localization (MADL) for location-dependent quantification of white matter hyperintensities. NeuroImage Clin. 2019;22:101772. doi:10.1016/j.nicl.2019.101772

30. Schirmer MD, Giese AK, Fotiadis P, et al. Spatial Signature of White Matter Hyperintensities in Stroke Patients. Front Neurol. 2019;10:208. doi:10.3389/fneur.2019.00208

31. Brickman AM, Zahodne LB, Guzman VA, et al. Reconsidering harbingers of dementia: progression of parietal lobe white matter hyperintensities predicts Alzheimer’s disease incidence. Neurobiol Aging. 2015;36(1):27–32. doi:10.1016/j.neurobiolaging.2014.07.019

32. Murray ME, Senjem ML, Petersen RC, et al. Functional impact of white matter hyperintensities in cognitively normal elderly subjects. Arch Neurol. 2010;67(11):1379–1385. doi:10.1001/archneurol.2010.280

33. Bunce D, Anstey KJ, Cherbuin N, et al. Cognitive deficits are associated with frontal and temporal lobe white matter lesions in middle-aged adults living in the community. PloS One. 2010;5(10):e13567. doi:10.1371/journal.pone.0013567

34. Tullberg M, Fletcher E, DeCarli C, et al. White matter lesions impair frontal lobe function regardless of their location. Neurology. 2004;63(2):246–253. doi:10.1212/01.wnl.0000130530.55104.b5

35. Ai Q, Pu YH, Sy C, Liu LP, Gao PY. Impact of regional white matter lesions on cognitive function in subcortical vascular cognitive impairment. Neurol Res. 2014;36(5):434–443. doi:10.1179/1743132814Y.0000000354

36. Auer DP, Pütz B, Gössl C, Elbel G, Gasser T, Dichgans M. Differential lesion patterns in CADASIL and sporadic subcortical arteriosclerotic encephalopathy: MR imaging study with statistical parametric group comparison. Radiology. 2001;218(2):443–451. doi:10.1148/radiology.218.2.r01fe24443

37. Brugulat-Serrat A, Salvadó G, Sudre CH, et al. Patterns of white matter hyperintensities associated with cognition in middle-aged cognitively healthy individuals. Brain Imaging Behav. 2020;14(5):2012–2023. doi:10.1007/s11682-019-00151-2

38. Fang E, Fartaria MJ, Ann CN, et al. Clinical correlates of white matter lesions in Parkinson’s disease using automated multi-modal segmentation measures. J Neurol Sci. 2021;427:117518. doi:10.1016/j.jns.2021.117518

39. McGuire S, Sherman P, Profenna L, et al. White matter hyperintensities on MRI in high-altitude U-2 pilots. Neurology. 2013;81(8):729–735. doi:10.1212/WNL.0b013e3182a1ab12

40. Rostrup E, Gouw AA, Vrenken H, et al. The spatial distribution of age-related white matter changes as a function of vascular risk factors--results from the LADIS study. NeuroImage. 2012;60(3):1597–1607. doi:10.1016/j.neuroimage.2012.01.106

41. Smith EE, Salat DH, Jeng J, et al. Correlations between MRI white matter lesion location and executive function and episodic memory. Neurology. 2011;76(17):1492–1499. doi:10.1212/WNL.0b013e318217e7c8

42. Al-Janabi OM, Brown CA, Bahrani AA, et al. Distinct White Matter Changes Associated with Cerebrospinal Fluid Amyloid-β1-42 and Hypertension. J Alzheimers Dis JAD. 2018;66(3):1095–1104. doi:10.3233/JAD-180663

43. Altermatt A, Gaetano L, Magon S, et al. Clinical associations of T2-weighted lesion load and lesion location in small vessel disease: Insights from a large prospective cohort study. NeuroImage. 2019;189:727–733. doi:10.1016/j.neuroimage.2019.01.052

44. Dalaker TO, Larsen JP, Dwyer MG, et al. White matter hyperintensities do not impact cognitive function in patients with newly diagnosed Parkinson’s disease. NeuroImage. 2009;47(4):2083–2089. doi:10.1016/j.neuroimage.2009.06.020

45. Damulina A, Pirpamer L, Seiler S, et al. White Matter Hyperintensities in Alzheimer’s Disease: A Lesion Probability Mapping Study. J Alzheimers Dis JAD. 2019;68(2):789–796. doi:10.3233/JAD-180982

46. Graff-Radford J, Arenaza-Urquijo EM, Knopman DS, et al. White matter hyperintensities: relationship to amyloid and tau burden. Brain J Neurol. 2019;142(8):2483–2491. doi:10.1093/brain/awz162

47. Holland CM, Smith EE, Csapo I, et al. Spatial distribution of white-matter hyperintensities in Alzheimer disease, cerebral amyloid angiopathy, and healthy aging. Stroke. 2008;39(4):1127–1133. doi:10.1161/STROKEAHA.107.497438

48. Jefferson AL, Holland CM, Tate DF, et al. Atlas-derived perfusion correlates of white matter hyperintensities in patients with reduced cardiac output. Neurobiol Aging. 2011;32(1):133–139. doi:10.1016/j.neurobiolaging.2009.01.011

49. Moscoso A, Rey-Bretal D, Silva-Rodríguez J, et al. White matter hyperintensities are associated with subthreshold amyloid accumulation. NeuroImage. 2020;218:116944. doi:10.1016/j.neuroimage.2020.116944

50. Quattrocchi CC, Errante Y, Mallio CA, et al. Inverse spatial distribution of brain metastases and white matter hyperintensities in advanced lung and non-lung cancer patients. J Neurooncol. 2014;120(2):321–330. doi:10.1007/s11060-014-1554-7

51. Torso M, Serra L, Giulietti G, et al. Strategic lesions in the anterior thalamic radiation and apathy in early Alzheimer’s disease. PloS One. 2015;10(5):e0124998. doi:10.1371/journal.pone.0124998

52. Saka E, Dogan EA, Topcuoglu MA, Senol U, Balkan S. Linear measures of temporal lobe atrophy on brain magnetic resonance imaging (MRI) but not visual rating of white matter changes can help discrimination of mild cognitive impairment (MCI) and Alzheimer’s disease (AD). Arch Gerontol Geriatr. 2007;44(2):141–151. doi:10.1016/j.archger.2006.04.006

53. Stenset V, Hofoss D, Berstad AE, Negaard A, Gjerstad L, Fladby T. White matter lesion subtypes and cognitive deficits in patients with memory impairment. Dement Geriatr Cogn Disord. 2008;26(5):424–431. doi:10.1159/000165355

54. De Groot JC, De Leeuw FE, Oudkerk M, et al. Periventricular cerebral white matter lesions predict rate of cognitive decline. Ann Neurol. 2002;52(3):335–341. doi:10.1002/ana.10294

55. Charidimou A, Boulouis G, Haley K, et al. White matter hyperintensity patterns in cerebral amyloid angiopathy and hypertensive arteriopathy. Neurology. 2016;86(6):505–511. doi:10.1212/WNL.0000000000002362

56. Griffanti L, Jenkinson M, Suri S, et al. Classification and characterization of periventricular and deep white matter hyperintensities on MRI: A study in older adults. NeuroImage. 2018;170:174–181. doi:10.1016/j.neuroimage.2017.03.024

57. Kim JH, Hwang KJ, Kim JH, Lee YH, Rhee HY, Park KC. Regional white matter hyperintensities in normal aging, single domain amnestic mild cognitive impairment, and mild Alzheimer’s disease. J Clin Neurosci Off J Neurosurg Soc Australas. 2011;18(8):1101–1106. doi:10.1016/j.jocn.2011.01.008

58. Bae S, Kim JE, Hwang J, et al. Increased prevalence of white matter hyperintensities in patients with panic disorder. J Psychopharmacol Oxf Engl. 2010;24(5):717–723. doi:10.1177/0269881108098476

59. Weaver NA, Doeven T, Barkhof F, et al. Cerebral amyloid burden is associated with white matter hyperintensity location in specific posterior white matter regions. Neurobiol Aging. 2019;84:225–234. doi:10.1016/j.neurobiolaging.2019.08.001

60. Biesbroek JM, Kuijf HJ, van der Graaf Y, et al. Association between subcortical vascular lesion location and cognition: a voxel-based and tract-based lesion-symptom mapping study. The SMART-MR study. PloS One. 2013;8(4):e60541. doi:10.1371/journal.pone.0060541

61. Yoshita M, Fletcher E, Harvey D, et al. Extent and distribution of white matter hyperintensities in normal aging, MCI, and AD. Neurology. 2006;67(12):2192–2198. doi:10.1212/01.wnl.0000249119.95747.1f

62. Gaubert M, Lange C, Garnier-Crussard A, et al. Topographic patterns of white matter hyperintensities are associated with multimodal neuroimaging biomarkers of Alzheimer’s disease. Alzheimers Res Ther. 2021;13(1):29. doi:10.1186/s13195-020-00759-3

63. Birdsill AC, Koscik RL, Jonaitis EM, et al. Regional white matter hyperintensities: aging, Alzheimer’s disease risk, and cognitive function. Neurobiol Aging. 2014;35(4):769–776. doi:10.1016/j.neurobiolaging.2013.10.072

64. Grey MT, Mitterová K, Gajdoš M, et al. Differential spatial distribution of white matter lesions in Parkinson’s and Alzheimer’s diseases and cognitive sequelae. J Neural Transm Vienna Austria 1996. 2022;129(8):1023–1030. doi:10.1007/s00702-022-02519-z

65. Phuah CL, Chen Y, Strain JF, et al. Association of Data-Driven White Matter Hyperintensity Spatial Signatures With Distinct Cerebral Small Vessel Disease Etiologies. Neurology. 2022;99(23):e2535–e2547. doi:10.1212/WNL.0000000000201186

66. Wen W, Sachdev P. The topography of white matter hyperintensities on brain MRI in healthy 60-to 64-year-old individuals. NeuroImage. 2004;22(1):144–154. doi:10.1016/j.neuroimage.2003.12.027

67. Wen W, Sachdev PS, Li JJ, Chen X, Anstey KJ. White matter hyperintensities in the forties: Their prevalence and topography in an epidemiological sample aged 44–48. Hum Brain Mapp. 2008;30(4):1155–1167. doi:10.1002/hbm.20586

68. Garnier-Crussard A, Bougacha S, Wirth M, et al. White matter hyperintensity topography in Alzheimer’s disease and links to cognition. Alzheimers Dement. 2022;18(3):422–433. doi:10.1002/alz.12410

69. Thu NT, Graff-Radford J, Machulda MM, et al. Regional white matter hyperintensities in posterior cortical atrophy and logopenic progressive aphasia. Neurobiol Aging. 2022;119:46–55. doi:10.1016/j.neurobiolaging.2022.07.008

70. Kamson DO, Illés Z, Aradi M, et al. Volumetric comparisons of supratentorial white matter hyperintensities on FLAIR MRI in patients with migraine and multiple sclerosis. J Clin Neurosci Off J Neurosurg Soc Australas. 2012;19(5):696–701. doi:10.1016/j.jocn.2011.07.044

71. Chou IJ, Lin KL, Wong AM, et al. Neuroimaging correlation with neurological severity in tuberous sclerosis complex. Eur J Paediatr Neurol EJPN Off J Eur Paediatr Neurol Soc. 2008;12(2):108–112. doi:10.1016/j.ejpn.2007.07.002

72. Schmidt P, Gaser C, Arsic M, et al. An automated tool for detection of FLAIR-hyperintense white-matter lesions in Multiple Sclerosis. NeuroImage. 2012;59(4):3774–3783. doi:10.1016/j.neuroimage.2011.11.032

73. Griffanti L, Zamboni G, Khan A, et al. BIANCA (Brain Intensity AbNormality Classification Algorithm): A new tool for automated segmentation of white matter hyperintensities. NeuroImage. 2016;141:191–205. doi:10.1016/j.neuroimage.2016.07.018

74. Bazin PL, Cuzzocreo JL, Yassa MA, et al. Volumetric neuroimage analysis extensions for the MIPAV software package. J Neurosci Methods. 2007;165(1):111–121. doi:10.1016/j.jneumeth.2007.05.024

75. Grimaud J, Lai M, Thorpe J, et al. Quantification of MRI lesion load in multiple sclerosis: a comparison of three computer-assisted techniques. Magn Reson Imaging. 1996;14(5):495–505. doi:10.1016/0730-725x(96)00018-5

76. van der Holst HM, van Uden IWM, Tuladhar AM, et al. Cerebral small vessel disease and incident parkinsonism: The RUN DMC study. Neurology. 2015;85(18):1569–1577. doi:10.1212/WNL.0000000000002082

77. Roseborough A, Ramirez J, Black SE, Edwards JD. Associations between amyloid β and white matter hyperintensities: A systematic review. Alzheimers Dement. 2017;13(10):1154–1167. doi:10.1016/j.jalz.2017.01.026

78. Schirmer MD, Bonkhoff A, Giese AK, Rost NS. Abstract WP179: Menopause Is Related To Accelerated Accumulation Of White Matter Hyperintensity Burden In Small Artery Occlusion Strokes. Stroke. 2023;54(Suppl_1):AWP179–AWP179. doi:10.1161/str.54.suppl_1.WP179

79. de Leeuw FE, de Groot JC, Achten E, et al. Prevalence of cerebral white matter lesions in elderly people: a population based magnetic resonance imaging study. The Rotterdam Scan Study. J Neurol Neurosurg Psychiatry. 2001;70(1):9–14. doi:10.1136/jnnp.70.1.9

80. van den Heuvel DMJ, Admiraal-Behloul F, ten Dam VH, et al. Different progression rates for deep white matter hyperintensities in elderly men and women. Neurology. 2004;63(9):1699–1701. doi:10.1212/01.wnl.0000143058.40388.44

